# nSIGHT™: A Data Discovery Platform for Visualization, Integration and Retrospective Analysis of Multimodal Clinical Research Data

**DOI:** 10.64898/2026.03.10.26347202

**Authors:** Mohammad Zia, Ben Plessinger, Kevin Eng, Aaron Flierl, Melissa Wilbert, Kelly Jans, Philip Whalen, Sarah Mullin, Joyce Ohm, Anurag K. Singh, Mark Farrugia, Carl Morrison, Christopher Darlak, Mukund Seshadri

## Abstract

The lack of interoperability among clinical and research data systems poses a significant barrier to cancer researchers interested in evaluating novel mechanistic hypotheses or translating innovative treatment strategies from the laboratory to the clinic. To address this gap in knowledge, we developed an innovative, web-based, data discovery, visualization and analysis tool (nSight™) that allows researchers to quickly and easily query clinical/research data and construct de-identified cancer cohorts. Guiding principles for development of the tool were focused on ease of use, intuitiveness, self-service, and presentation of structured but de-identified data to the end user. nSight™ provides users with information on patient demographics, disease histology, diagnostic procedures and therapeutic interventions, timeline of disease progression/recurrence, along with available molecular profiling/sequencing data and indicators of participation in epidemiologic or lifestyle studies for specific cancer patient cohorts. The platform also allows users to obtain summary statistics based on demographic, histologic and clinical factors as well as perform basic survival analysis using Kaplan-Meier curves between specific patient cohorts. nSight™ is an intuitive, user-friendly tool that enables visualization, integration and analysis of multimodal clinical and research data without placing high technical demands or time constraints on researchers. The platform is designed for research feasibility assessment, cohort development, and retrospective data discovery, which in turn should help investigators identify potential study populations and explore novel hypotheses.

## INTRODUCTION

Comprehensive cancer centers often require the establishment and maintenance of multiple disparate information technology systems and data warehouses to meet their clinical, administrative/financial, regulatory and research needs. These systems often operate in silos and are typically designed to meet specific needs with limited standardization on data architecture or interoperability (1). For example, metadata on patient/tumor characteristics and treatment outcomes are housed in secure clinical data systems (e.g. electronic medical records, laboratory information management systems) to ensure regulatory compliance and maximize patient safety/privacy. In comparison, large amounts of sequencing data are typically stored in research data systems within institutional biorepositories or shared resources that do not communicate or interface with clinical data systems. Extracting meaningful information from these disparate resources is laborious, time-consuming, and often requires domain expertise in informatics or programming (2). The lack of interoperability among clinical and research data systems combined with the need for specialized expertise is a major barrier for basic scientists interested in translating innovative discoveries or treatment strategies from the laboratory bench to the clinic (2, 3).

To address this challenge, we developed an innovative, web-based, data discovery tool (nSight™) that allows researchers to quickly and easily query clinical and research data to construct de-identified cancer cohorts. To facilitate its adoption by the broad research community, guiding principles for its development were focused on ease of use, intuitiveness, self-service, and representation of the data in a de-identified state to enable unrestricted “fast fail” exploration of hypotheses. The platform was designed for research feasibility assessment, cohort development, and retrospective data discovery to help investigators identify potential study populations and explore novel hypotheses. Specifically, nSight™ provides users with a timeline of oncology care events, including diagnosis, start and completion of treatment, disease progression/recurrence, along with tumor molecular profiling data for individual patients and cohorts. The platform also allows users to obtain summary statistics based on demographic, histologic and clinical factors as well as perform basic survival analysis using Kaplan Meier Curves, and p-value calculations to establish difference between cohorts. Here, we report on the development of the system architecture and technical implementation of this platform along with its deployment and demonstration of its utility as a data exploration tool in head and neck cancer.

## MATERIALS AND METHODS

### Data sources, Extraction and Transformation

The searchable database in nSight™ was populated from data extracted, transformed, and combined from nine different data sources: cancer registry, electronic health record (EHR), laboratory information management system (LIMS), radiology information system (RIS), patient surgery management software, Patient Registration System, clinical trial management system, IRB management system, and electronic medical records (EMR). In addition, raw files from molecular diagnostic laboratories were also parsed to populate the database. Data from all these sources were transformed and combined into 21 event types, and 6 different event categories in Python (Version 3.13). An event contained an information-rich label along with relative number of days, months, and years since the date of diagnosis and the percent of time elapsed between date of diagnosis and last contact date or death date. Along with transforming the source data into events, searchable terms were extracted from some of the events. These terms were used in the search omni bar to enable searching. Demographic information for the patients was also extracted from cancer registry, EHR, and the patient registration system.

### Standardization

Integrating data from different sources requires interoperability and standardization of source terminologies (4, 5). Given the widespread adoption and the introduction of an oncology module, we have adopted OHDSI’s standardized vocabularies for multiple event domains: Patient and Therapeutic (Chemical and Procedure). This includes mapping ICD-10-CM and ICD-O-3 to SNOMED CT (Systematized Nomenclature of MEDicine Clinical Terms) (6) and national drug codes (NDC) (7) and proprietary drug names to RxNorm. The details for each event type and their searchable terms are described below.

### Disease Events

#### Diagnosis

Anchor event that provided the date of diagnosis and all other event types were reported relative to this event. Using an anchoring event for relative timelines, rather than specific dates, enhanced de-identification of the product. The source for this diagnosis event was cancer registry. The label for this event contained age at diagnosis, primary site, histology, clinical grading, and pathological grading information. The searchable terms for this event were primary site, primary site group, histology, histology group, clinical grading, and pathological grading.

#### First Recurrence

This event indicated the time of recurrence as reported in the cancer registry. The label for this event contained information about the type of recurrence. The searchable terms for this event were the recurrence type only.

#### Disease/Patient Status

This event indicated if the patient was alive or deceased based on last date of contact or when they were reported as deceased. The source for this event was cancer registry and EHR. The searchable terms for this event were alive or deceased status.

### Patient Events

#### Medical history

This event indicated ICD-10 and SNOMED diagnosis codes for the patient as reported in the EHR. When possible, SNOMED codes were converted to ICD-10 codes. The event label contained the full ICD-10/SNOMED code and description. The searchable terms were the code and description. This event indicated ICD-10 and SNOMED diagnosis codes for the patient’s family history as reported in the EHR. When possible, SNOMED codes were converted to ICD-10 codes. The event label contained the full ICD-10/SNOMED code and description. The searchable terms were the code and description.

### Therapeutic Interventions

#### Prescribed and administered drugs

This event indicated drugs that were prescribed, including historical (self-reported), prescribed and administered drugs. The source for this data was the EHR. All brand name drugs were converted to generic names. The label for this event contained the generic name, therapeutic category, and the parent therapeutic category of the drug. The searchable terms were the generic drug name, therapeutic category, and parent therapeutic category.

#### Surgery

This event indicated description of any surgery at the institute or outside the institute. The sources for this data were the EHR, LIMS, and patient surgery management software. The label for this event was the description of the surgery as entered into the source system.

#### Radiation Therapy

This event indicated when radiation therapy was started. The source for this data was the EMR. The label for this event included the site of radiation, type, and dosage. The searchable term was the radiation type.

#### Clinical Trial

This event indicated when a patient started treatment on a clinical trial, when the treatment ended, and when they were taken off the clinical trial. The source for this data was the institute’s clinical trial management system. The label for this event contained the trial identifier, principal investigator, and description.

### Biospecimen Events

#### Frozen Tissue

This event indicated the presence of frozen tissue from a given patient. The source for this data was LIMS. The event label contained the frozen tissue site, histology, and quantity.

#### Formalin-fixed, paraffin-embedded (FFPE) tissue

This event indicated the presence of FFPE tissues in institute biorepositories. The sources for this data were LIMS and cancer registry since the latter could indicate potential FFPEs based on surgery information. The event label contained the surgical event ID, site, and histology if the source is LIMS; otherwise, it contained the surgery description with a disclaimer about it being a potential FFPE.

#### BioRepository and laboratory services (BLS)

This event indicated the presence of blood samples. The source for this data was LIMS. The event label merely contained the collection ID.

#### Tissue Microarray (TMA)

This event indicated the presence of TMAs. The source for this data was LIMS. The event label contained tissue type, TMA description, and TMA ID.

#### Hematology Collection

This event indicated the presence of hematology collection. The source for this data was LIMS. The event label contained the collection ID and quantity.

#### Patient-Derived Xenograft (PDX) Models

This event indicated the presence of PDX models. The data was sourced from LIMS and the event label contained the tissue site, PDX model ID, gender, and passage information.

### Diagnostic Events

#### Clinical Genomics

This event indicated the presence of results from molecular diagnostic laboratories. The sources for this data were XML, CSV, and JSON files sent back from the laboratories. Because the laboratories reported the results differently, significant harmonization was required. The event label contained the laboratory name, the diagnostic test performed, gene, variant, cancer mutation hotspot, mutation burden score, and microsatellite instability status. The searchable terms were gene, variant, if mutation burden score was greater than 10, and if the microsatellite instability status was high.

#### Single Analyte Test

This event indicated the presence of results from a molecular diagnostic laboratory. The source for this data was CSV files sent back from a laboratory. The event label included the test performed and results. The searchable terms were the test performed and results.

### Research Data

#### Research Sequencing

This event indicated the presence of any sequencing data (DNA, RNA) available on the patient tumors. The source data was obtained from LIMS. The event label contained the request ID, requesting Principal Investigator (PI), sequencing type, and library type.

#### Biorepository Questionnaire

This event indicated the presence of Biorepository & Laboratory Services (BLS) survey or questionnaire data. The source data was obtained from LIMS. The event label indicated the questionnaire ID.

### Data Visualization and Analysis

A patient-specific timeline view of all medical events was created in D3.js (v 7.6.1). Pie charts and bar graphs were written in Chart.js (v.4.0.1 ng2-charts) to summarize search results. Kaplan-Meier survival analysis is performed using the LIFELINES Python package and all p-values were calculated using the Scikit Python package.

## RESULTS

### Design and development of nSight™ architecture

The overall framework and the extract, transform, and combine (ETC) process used to design and develop nSight*™* is shown schematically in **Figure 1**. Raw data was extracted from multiple clinical sources **(Fig. 1A)** using multiprocessing Python scripts that ran in parallel and stored the extracted data into SQLite3 databases (**Fig. S1**). The ETC steps were performed on a virtual machine (VM) server that was authorized to contain PHI (**Supplementary Methods and Supplementary Figure 1**). This VM was deployed within the internal network at Roswell Park and access required a VPN or a direct connection to the cancer center’s internal network, with SSH permissions strictly limited to members of the design team.

**Figure 1.**
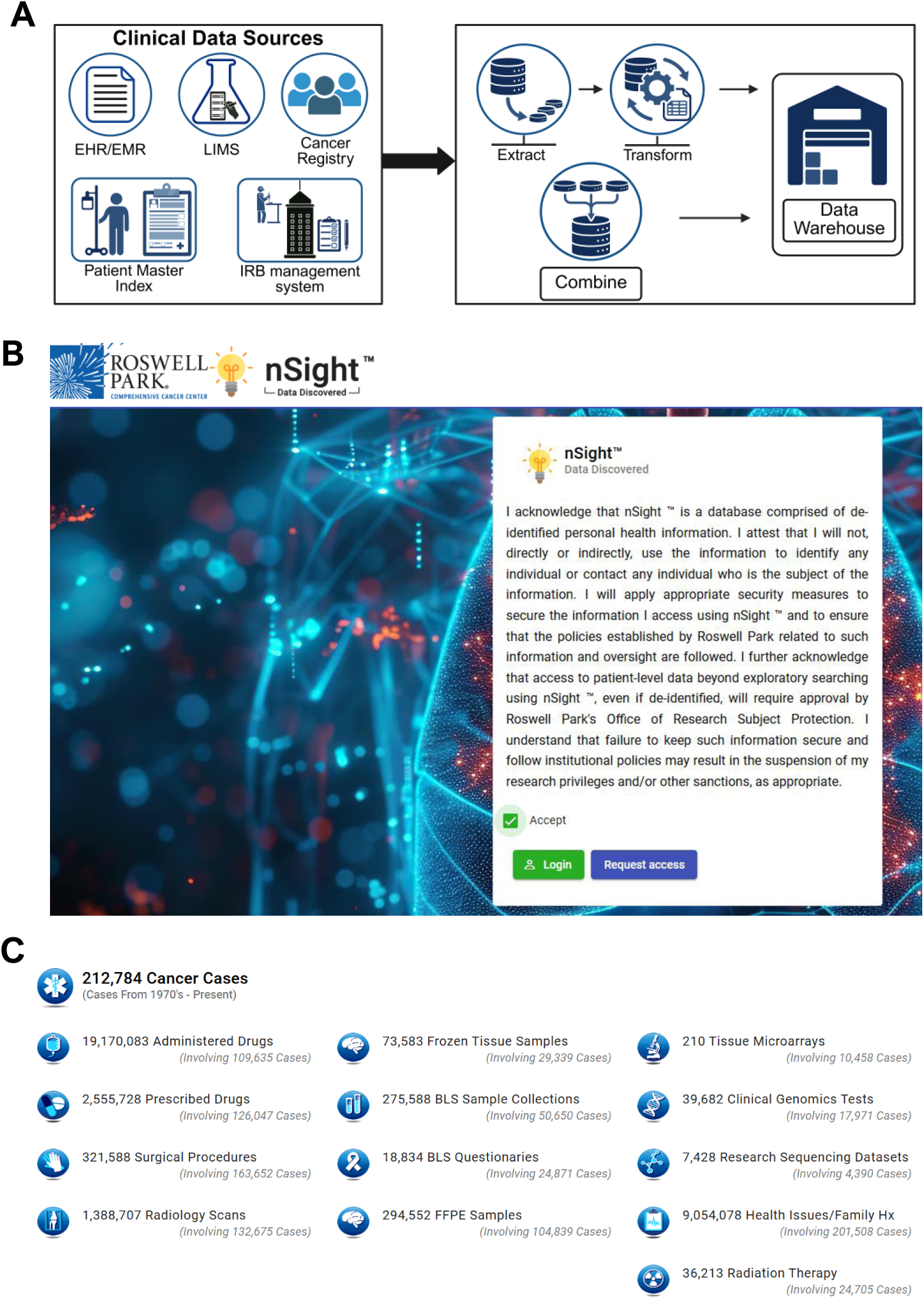
Design framework and user interface of nSight™. (**A**) The extract, transform, and combine (ETC) process used to design and develop nSight™. Raw data was extracted from multiple clinical sources and stored in SQLite3 databases. The “extracted” databases were subsequently transformed into events relative to the date of diagnosis and searchable terms were extracted and stored into SQLite3 “transformed” databases. The transformed databases were combined to create a single SQLite3 combined database. (**B**) A web-based interface allows users to search for cancer case histories and display associated events on a timeline for each patient. Approved users are first required to acknowledge and comply with all institutional policies and apply appropriate security measures to information accessed using nSight™. (**C**) Snapshot of the available cancer cases, diagnostic, research, and treatment information available to authorized users of nSight™.

### nSight™ user interface

A web-based interface was written in Angular with supporting REST API in Python/Django to allow users to search for cancer case histories and display the associated events on a timeline for each patient. As shown in **Figure 1B**, approved users are first required to acknowledge and comply with all institutional policies and apply appropriate security measures to information accessed using nSight™. A snapshot of the multimodal dataset available to users (Accessed on Oct 10, 2025) is shown in **Figure 1C**. Once logged onto the platform, users are required to create a workspace, specifying a name, description and an optional IRB-protocol number (**Fig**. **2A**). As shown in **Figure 2B**, the interface allows users to identify case histories based on gender, patient status, and presence of events through a quick search box. Additionally, three omni search bars allow users to search for events based on specific terms under inclusion and exclusion criteria, as applicable. The search function can be performed with all search terms included or be matched to one or more terms.

**Figure 2.**
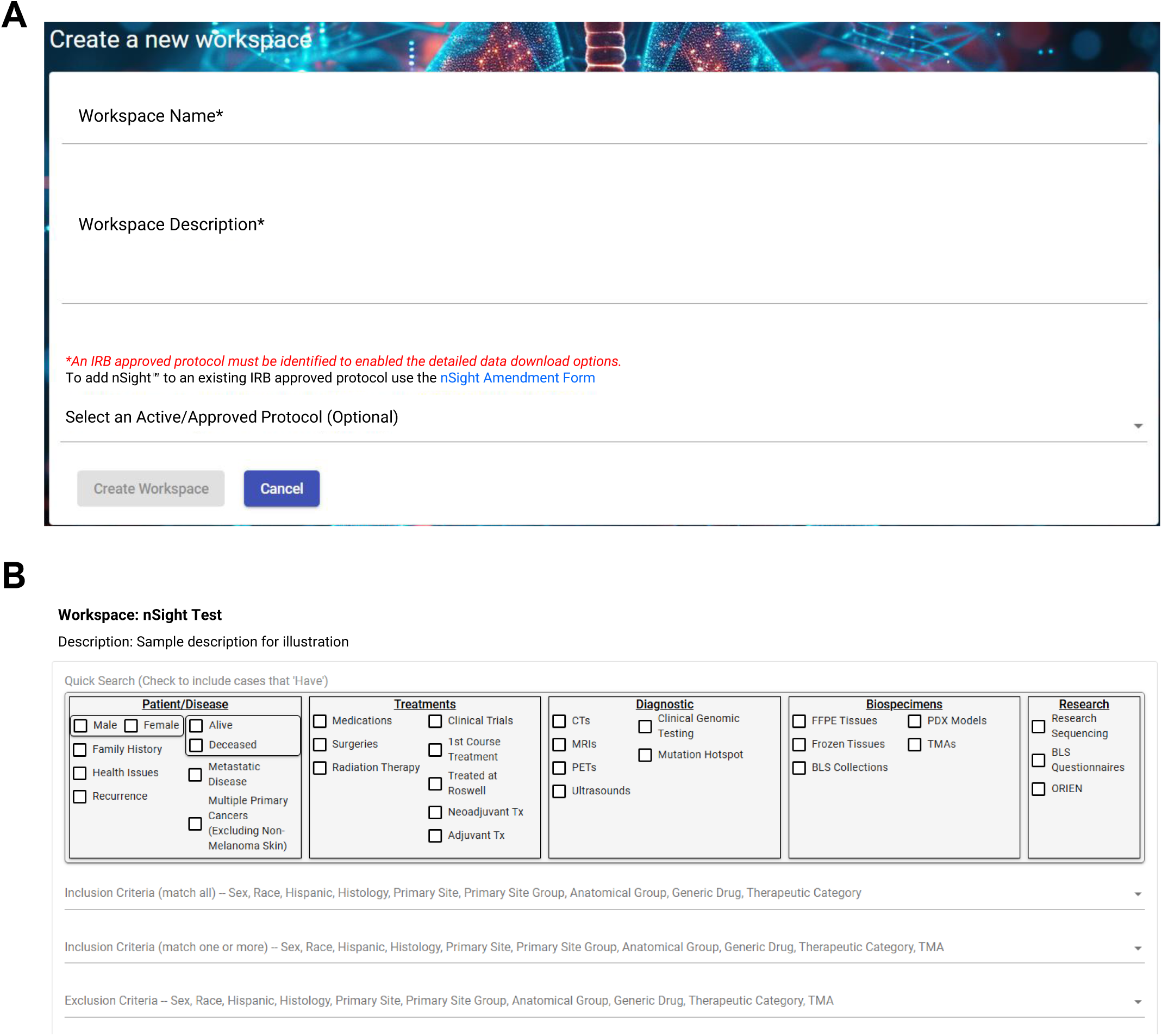
nSight™ User Workspace and Search Terms. (**A**) Approved nSight™ users are required to create a workspace with basic information. Creation of the workspace allows users to create specific cohorts of patients based on probing for a specific cancer type and treatment, or based on available diagnostic tests, biospecimens, and sequencing data. (**B**) The interface allows users to identify case histories based on gender, patient status, and presence of events through a quick search box. Additionally, three omni search bars allow users to search for events based on specific terms (described under the Materials and Methods section) under inclusion and exclusion criteria.

### Timeline of patient-specific clinical events using nSight™

Creation of the workspace allows users to create specific cohorts of patients based on a specific cancer type and treatment, or based on available diagnostic tests, biospecimens, and sequencing data. To enable easy visualization of history and all associated medical events for specific cancer cohorts, a patient-specific timeline view was created, with the x-axis used to display the time scale since diagnosis (in days, months, years since diagnosis). The timeline can also be viewed as a relative measure of time elapsed between diagnosis and date of last contact or death, computed during the transformation. The illustration in **Figure 3** shows the timeline view for four distinct patients. The left side of the timeline contains PT-ID, age at diagnosis, sex, primary site of cancer, survival in number of days, green for alive and red for deceased, ethnicity status, and if the patient was treated at Roswell Park, indicated using the cancer center logo. Each event type is vertically stacked and for a given event type, the events are spread across the x-axis based on the timescale. The y-axis was used to stack different event types vertically, with each event type designated using a symbol and color. Hovering over the events displays the labels that were previously created during transformation. For a given event type, if duplicate labels were present, they were shown only once, but the number of times they occurred was indicated. The legend of the event types is shown on the right in **Figure 3**. A dropdown menu allows users to sort the timeline by PT-ID, sex, age at diagnosis, survival in number of days, patient status and change the x-axis scale. Users also have the ability to filter events based on the timeline implemented.

**Figure 3.**
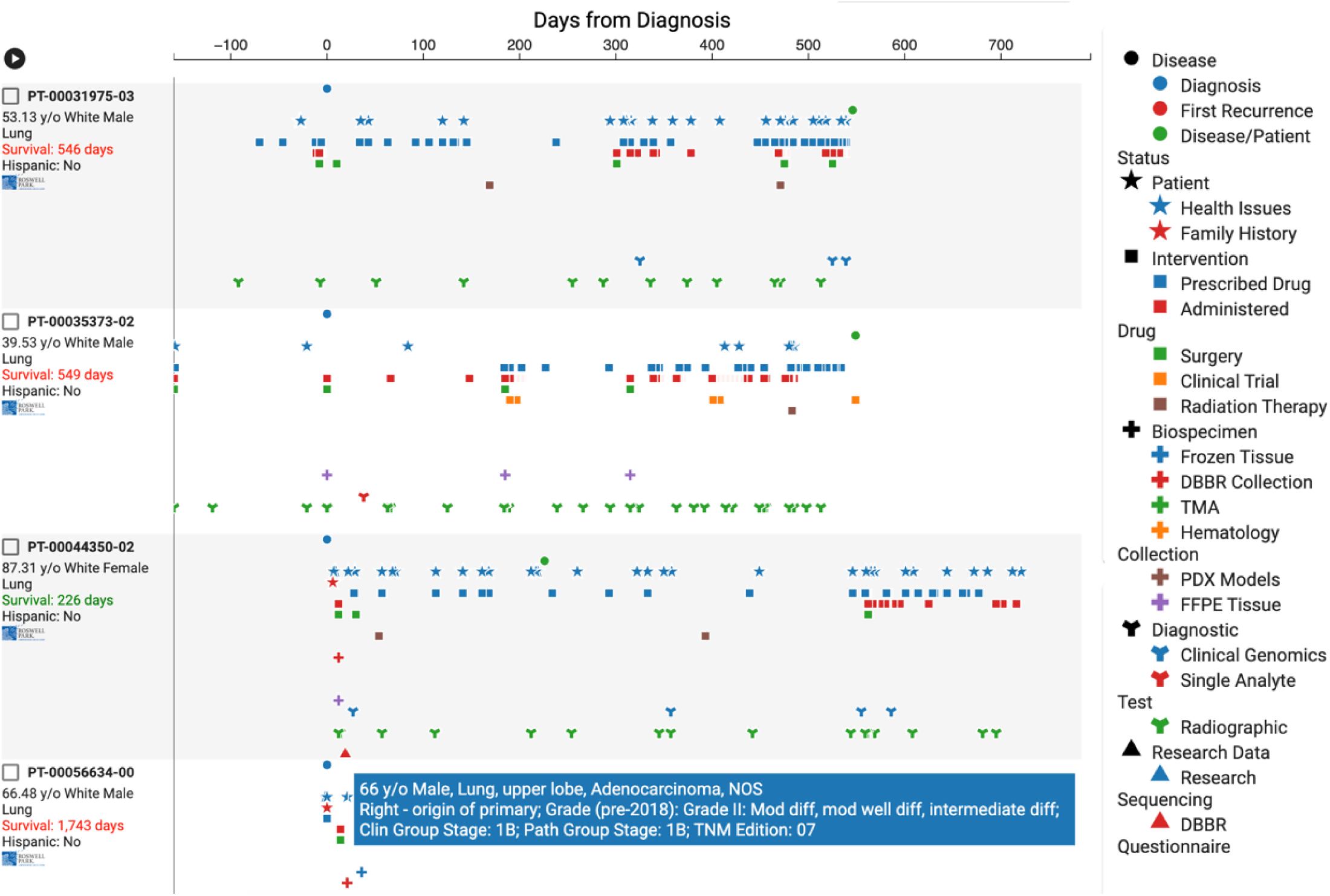
Timeline view of patient-specific events in nSight™. To enable easy visualization of history and all associated medical events for specific cancer cohorts, a patient-specific timeline view was created, with the x-axis used to display the time scale since diagnosis (in days, months, years since diagnosis). The timeline can also be viewed as a relative measure of time elapsed between diagnosis and date of last contact or death. Each event type is vertically stacked and designated by symbols. For a given event type, the events are spread across the x-axis based on the timescale.

### Data visualization and statistical capabilities of nSight™

The principal motivation behind development of nSight™ was to provide researchers with a user-friendly tool that enabled visualization of curated data extracted from a plurality of clinical and research data sources. To accomplish this, a ‘*Summary View*’ was created to display the results of a specific search by the user. This summary view includes information pertaining to patient demographics, disease, intervention, biospecimen, and the available research data from patients and/or patient tumors. The pie charts and bar graphs shown in **Figure 4** illustrate the wealth of information captured and stratified by nSight™ that is made available to users under separate tabs. The data summarized in **Figure 4A** pertains to information on patient demographics, including sex, race/ethnicity, alcohol/tobacco use and age at diagnosis. A sample summary view of disease-specific information including clinical and pathologic staging, primary cancer site, and histology is shown in **Figure 4B**. The bar graphs shown in **Figure 4C** illustrate sample data available to users on specific interventions including standard of care therapies, along with administered drug groups. Additionally, information on participation in interventional clinical trials is also available. The data visualization tool also allows users to obtain information on the availability of biospecimens (**Fig. 5A**) from the Biorepository & Laboratory Services (BLS) collection at Roswell Park, including fresh and frozen tissues, formalin fixed paraffin embedded (FFPE) samples, tissue microarrays (TMAs), and patient-derived xenograft (PDX) models generated from donor human tumor tissues. And finally, users can review the sequencing data (**Fig. 5B**) available for samples within a given cancer cohort based on their selection criteria.

**Figure 4.**
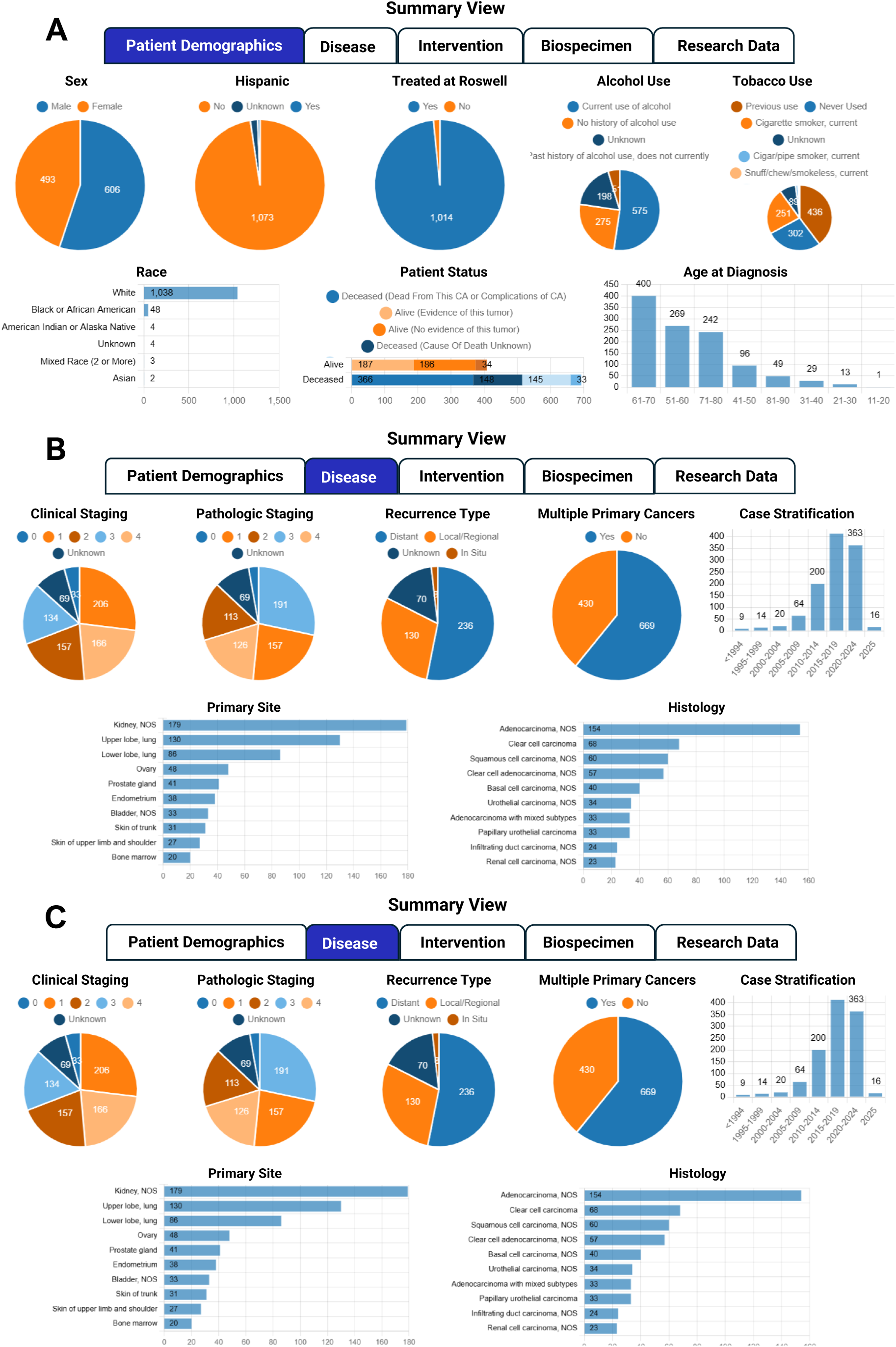
Summary view of clinical data on nSight™. The ‘Summary View’ within nSight™ displays the results of a specific search by the user. This includes information on patient demographics (**A**), disease-specific information (**B**), treatment information including standard of care therapies, administered drugs and clinical trial participation by patients (**C**). This allows authorized users to visualize curated data extracted from a plurality of data sources.

**Figure 5.**
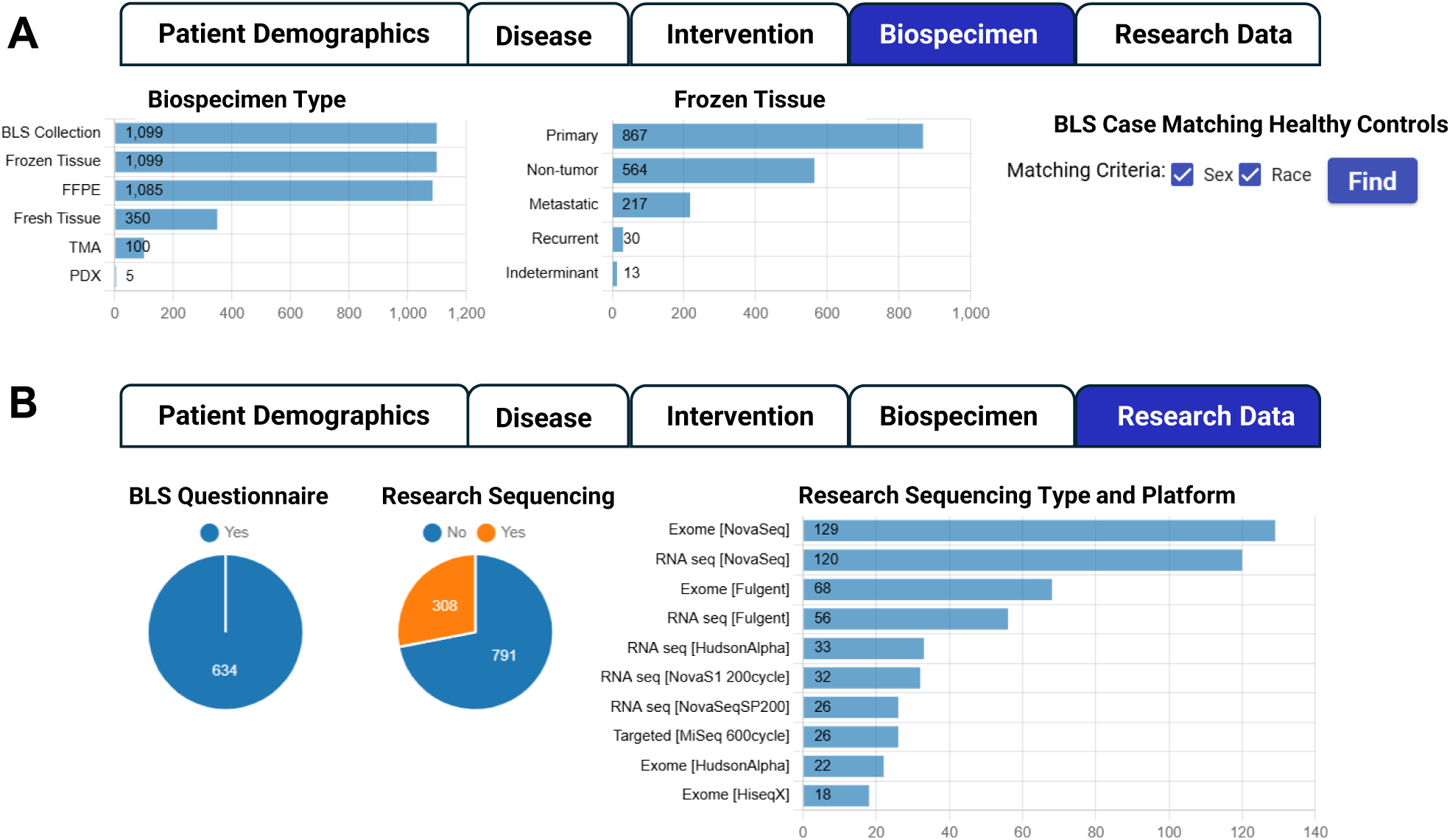
Summary view of research data on nSight™. The ‘Summary View’ within nSight™ displays the results of a specific search by the user. This includes information on biospecimens collected as part of clinical research studies under approved protocols along with patient responses to questionnaires collected as a part of epidemiologic or biobehavioral studies (**A**), and available sequencing data and platform (**B**).

And finally, to allow users to perform comparisons between two cancer cohorts or subsets of patients, an ‘Output’ tab was created that can combine case histories from multiple groups and organize them into a table. This option also enables users to perform basic statistical analyses. For continuous variables, the Kruskal-Wallis H-test is used, while the Chi-square test is used for comparing categorical values and calculating p-values. The tool can also perform basic Kaplan-Meier survival curve(s) and median survival times based on log rank test. Users can download the characteristics table, the case history IDs, and case history details as an Excel spreadsheet. Users with an approved IRB-approved protocol can export all the event data in a table format along with summary table as generated in the ‘Output’ tab.

### Training and Usage Results

One quantifiable way to measure adoption of any software platform is within the analysis of system usage over time. To educate users on the functionality and proper usage of nSight*™*, short 3–5-minute educational videos were produced and strategically embedded throughout the application. Written documentation was also made available on the website. Lastly, in-person workshops were held to provide hands-on training on nSight™. The training was conducted in collaboration with ‘honest brokers’ at the cancer center. A Microsoft Power BI dashboard was created that provided usage statistics based on user to track the usage of nSight*™*. Utilizing eight months of data from nSight™ system usage logs, an initial baseline search per month shows 313 individual queries during the initial month of go-live. A significant increase in searching occurred in months 7-8 with a peak search per month of 465 individual queries run at month eight. Factors influencing system usage/monthly search rate include but are not limited to initial go-live curiosity/interest, grant/major meeting deadlines, timing of product marketing efforts, research disease group presentations, addition of new data sources as well as new feature releases. To date, the application has been utilized by over 100 individual members.

### Use-case example

To illustrate the utility of nSight™, we performed a simple evaluation of survival outcomes in patients with head and neck squamous cell carcinoma (HNSCC). Previous studies have highlighted older age as a prognostic factor that can predict oncologic outcomes in HNSCC patients undergoing chemoradiation (8, 9). To understand the impact of age on treatment outcomes in this patient population, we utilized nSight™ to identify cohorts and performed an exploratory analysis of all HNSCC patient data available through nSight™. First, all patients with a diagnosis of HNSCC were identified and information on patient demographics obtained (**Fig. 6 A-C**). Subsequently, the “Output” tab of nSight™ was used to combine case histories from multiple groups and stratify the data into two groups – Age 18-64 (**Fig. 6 D-F**) and patients over the age of 65 (**Fig. 6 G-I**). The built-in statistics tool was then used to compare differences in median survival times between the two groups using a log rank test. Kaplan-Meier curves (**Fig. 6J, K**) illustrate the significant differences in 5- and 10-year survival between the two cohorts. These findings were independently validated using a REDCap database that is maintained by the Department of Radiation Medicine at Roswell Park. Notably, this entire process for creating a workspace, querying for the specific cancer patient population, stratifying groups and performing the analysis using nSight™ took approximately 20 minutes, illustrating the utility of nSight™ as a rapid data discovery platform.

**Figure 6.**
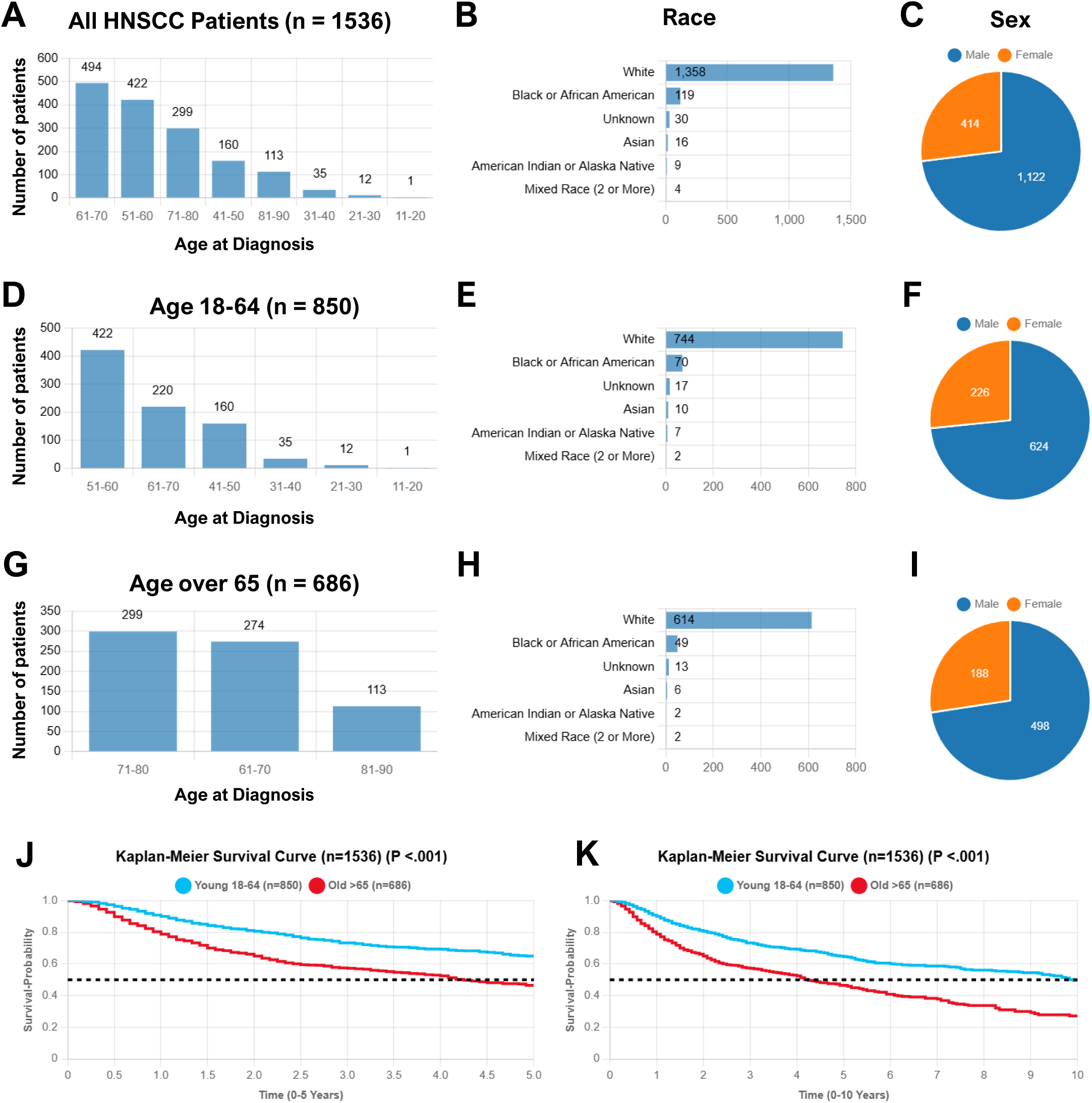
Sample subset comparisons and statistical analysis using nSight™. To demonstrate the utility of nSight™, patients with a diagnosis of head and neck squamous cell carcinoma (HNSCC) were identified (**A-C**). The “Output” tab of nSight™ was used to combine case histories from multiple groups and stratify the data into two groups based on age (**D-I**). The built-in statistics tool allowed for comparison of differences in median survival times between the two groups using on log rank test. Kaplan-Meier curves (**J, K**) illustrate the significant differences in 5- and 10-year survival between the two cohorts.

## DISCUSSION

Data harmonization is a recognized challenge in cancer research since clinical and research data are often stored in distinct systems that lack interoperability, due to regulatory and ethical requirements (1–3). Additionally, manual integration of large multimodal datasets to answer research questions is tedious and cost prohibitive. Development and validation of data discovery tools that can safely and reliably integrate multimodal clinical and research data is therefore critical to enable the conduct of clinical research studies in a time-efficient, and cost-effective manner. Here, we report on the design, development and utility of nSight™, an intuitive, user-friendly tool that enables visualization, integration and analysis of multimodal clinical and research data. The tool enables the conduct of clinical research studies without placing high technical demands or time constraints on researchers.

A significant amount of effort has gone into building data science infrastructure (e.g. Cancer Research Data Commons) to share the wealth of existing clinical cancer data with the research community and to leverage available electronic health information for secondary uses in clinical or translational cancer research (10, 11). Indeed, academic medical centers have also developed institutional resources and data discovery tools such as STARR (12), Leaf (13), POSEIDON (14), and MCAP (15). While the overall motivation for creating nSight™ was similar to these tools, several aspects of the design and framework of nSight™ are unique and noteworthy. The architecture of nSight™ was designed by the need to ensure compliance with all ethical and regulatory standards. First, visualization of all clinical data including timeline/episodes of care in a de-identified manner. To meet this requirement, a time point was selected as an anchor so that all other episodes of care could be reported relative to this anchor point. We chose the date of diagnosis as the anchor point and calculated the relative days, months, and years from the date of diagnosis for all the episodes of care. This was accomplished by using data from the cancer registry as the basis for defining events. This approach could therefore be replicated by other institutions with a clinical data warehouse similar to the cancer registry. The modular design of nSight™ allows for easy inclusion of new data sources. Second, to eliminate the need to store any PHI on the application server, we created an architecture composed of two servers. We extracted and transformed all the data on a dedicated extraction-transformation server whose access control we limited to a few individuals to avoid any PHI leak. We scheduled a nightly task to extract, transform, and transfer data to the application server, where we loaded the transformed data into Elasticsearch using another scheduled task. And finally, restricted access to authorized users who had completed all research training requirements and were approved to access associate clinical data under an approved protocol. To fulfill this requirement, we integrated our institute’s Azure Active Directory SSO and leveraged its group feature to limit access to nSight™. The management of nSight™’s Azure Active Directory group was also designated to institutional ‘honest brokers’, who reviewed all requests for data access. Only de-identified data was made available to end users of nSight™. The development of nSight™ was also conducted in accordance with institutional policies and under an institutional review board (IRB)-approved protocol at Roswell Park Comprehensive Cancer Center. Under the IRB-approved protocol, the development team was granted access to the data sources and all members of the development and testing team were required to complete Collaborative Institutional Training Initiative (CITI) modules on Human Subjects Research, including Biomedical Responsible Conduct of Research. Similarly, end users of the developed tool were required to complete CITI training modules prior to being given access to nSight^™^. Such data governance considerations should be taken into account by institutions while developing such informatic tools.

nSight™ leverages the Observational Medical Outcomes Partnership (OMOP) Common Data Model (CDM) ensuring data standardization across clinical datasets utilizing different terminological and semantic structures (4–7). Adoption of this widely used common data model standardization enables data sharing across institutions and allows researchers and data scientists to track mapping progress, assess data quality, and identify potential data gaps, reinforcing data integrity and analytical rigor. The OMOP CDM stewarded by the OHDSI community, provides a standardized structure and vocabulary for observational health data. Mapping of nSight™ to OMOP standard descriptions provides the ability to collaborate and compare data from other institutions that have also adopted the OMOP standard. Conversely, those who have adopted the OMOP CDM can easily configure their database to use nSight™ at their own institution. Additionally, when cohort data is downloaded from the ‘Output’ tab, the descriptions are predominantly OMOP standard descriptions where applicable, which directly enables researchers to compare data effectively.

The nSight™ data discovery tool utilizes a novel visualization technique allowing users to view an entire patient episode of care in a de-identified manner, visually marking important events (i.e., diagnosis, treatment, biospecimen collection etc.). Many interactive features within the visualization tool aid the user in finding specific cases of interest in line with their research objectives. These features include an ability to hover over symbols to gain specific knowledge of an event (e.g., tumor histology, stage, grade within diagnosis), as well as an ability to filter and/or highlight text within specific events (e.g., filter health issues where ‘nausea’ is present etc.). Other features include an ability to zoom in and out as well as panning left and right while exploring a patient’s episode of care. Another important solution for the discovery system is the ability for the user to save specific records into custom groups, allowing the user to create complex cohorts, and add sub-cohorts with labels and records of their choosing. The platform also allows users to perform basic statistical analyses including Kaplan Meier survival, and p-value calculations in real-time to determine statistical significance between observed differences between two cohorts.

It is important to recognize certain limitations of nSight™. One limitation of the underlying data model is the lack of relationships between many record types and the specific instance of disease. Except for data extracted from the organizational cancer registry system, most other data objects link at the patient level rather than to a tumor or disease instance. Some of this complexity is mitigated in the visualization, where data points are positioned relative to the diagnosis time point; however, many complex cases (e.g., multiple disease instances) still present challenges in serving data to researchers in a clear and comprehensible way. Another limitation is that cohorts are constructed using disease information. nSight™ is currently not equipped to include diagnostic imaging datasets (e.g., CT or MRI) and therefore does not allow users to integrate longitudinal radiologic data into their cohorts. With increasing interest in the application of artificial intelligence (AI) based approaches to imaging (‘radiomics’) (16) and multimodal data (‘radiogenomics’) (17), it would be useful to expand the utility of nSight™ to enable researchers to build curated collections of imaging data, filtered by patient cohort, and imaging modality. The utility of nSight™ could be further strengthened by integration of multimodal data with AI-based multiomic (radiomic, pathomic, genomic) analysis pipelines for diagnostic or prognostic applications (18).

In conclusion, the nSight™ system developed at Roswell Park Comprehensive Cancer Center is a valuable resource that allows researchers to evaluate research feasibility (based on cohort discovery), hypothesis development, and facilitates collaborative research through exchange of de-identified data. The tool allows researchers to create complex cohorts based on specific cancer types, genetic mutations, treatment modalities, and demographic factors, and supports further data exploration. The longitudinal data provided by nSight™ also offers the potential for investigators to study the evolution of specific cancer types as a function of the genetic profile of the patient/tumor, and in response to therapy. The creation of nSight™ adds to the limited number of discovery tools (e.g., STARR, Leaf) available to investigators for cohort discovery and to query/identify patients with specific clinical characteristics or exposures (treatments).

## Data and code availability statement

All data needed to evaluate the conclusions are presented in the paper. Raw data and codes can be made available upon reasonable request.

## Consent for publication

Not applicable

