## Supplementary Information for "nSIGHT™: A Data Discovery Platform for Visualization, Integration and Retrospective Analysis of Multimodal Clinical Research Data"

**SUPPLEMENTARY METHODS**

**ETC Framework**

The “extracted” databases contained the patient medical record number (MRN), and a text field containing all fields extracted from the source as a JSON string. These databases were subsequently transformed into events relative to the date of diagnosis and searchable terms were extracted and stored into SQLite3 “transformed” databases. These databases contained the institute’s participant ID (PT-ID), removing any identifying information by mapping MRN to PT-ID, and a JSON string with the events. The transformed databases did not contain any PHI, including MRNs or dates. The transformed databases were combined to create a single SQLite3 “combined” database. This database contained the PT-ID and a JSON string containing all the events and all searchable terms.

**ETL server configuration**

The ETL server was a Red Hat Enterprise Linux 9.6 (Plow) virtual machine configured with 8 vCPUs and 32GB of RAM. It hosts the nSight™ application, which is deployed using multiple containers, including an Nginx web server, a Python/Django backend, a MariaDB database, and three Elasticsearch instances. Elasticsearch (v8.5.0) was used to store and search the data transferred from the ETC VM server. A Python script was scheduled to run nightly to load the transferred data into Elasticsearch. Django-Backend (v4.1.3) powered the Django REST API (v3.14). The REST API managed the communication with the Angular Frontend and Django Q-Cluster. Angular-Frontend (v15.0.0) powered the user web interface. The angular application was served by Nginx (v1.23.2). Django-Q-Cluster (v1.3.6) handled asynchronous processing of data extraction requests, while MariaDB (v10.10.2) supported the Django application. Approved users with access to nSight*™* were required to authenticate using Azure Active Directory Single Sign-on (SSO).

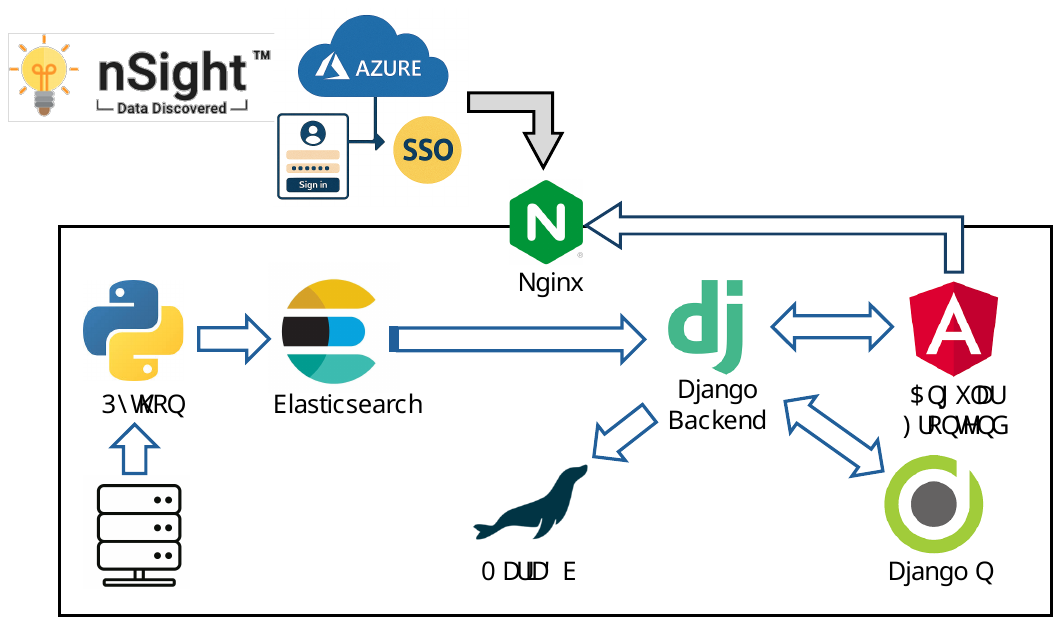

**Figure S1. Architecture of nSight™**. The extract, transform, and combine (ETC) process used to design and develop nSight™. The ETC steps were performed on a VM server that was authorized to contain PHI by the institute’s IT security. The ETC steps were scheduled to run nightly via cron job.
